# A one-year hospital-based prospective COVID-19 open-cohort in the Eastern Mediterranean region: The Khorshid COVID Cohort (KCC) study

**DOI:** 10.1101/2020.05.11.20096727

**Authors:** Ramin Sami, Forogh Soltaninejad, Babak Amra, Zohreh Naderi, Shaghayegh Haghjooy Javanmard, Bijan Iraj, Somayeh Haji Ahmadi, Azin Shayganfar, Mehrnegar Dehghan, Nilufar Khademi, Nastaran Sadat Hosseini, Mojgan Mortazavi, Marjan Mansourian, Miquel Angel Mañanas, Hamid Reza Marateb, Peyman Adibi

## Abstract

The COVID-19 is rapidly scattering worldwide, and the number of cases in the Eastern Mediterranean Region is rising. Thus, there is a need for immediate targeted actions. We designed a longitudinal study in a hot outbreak zone to analyze the serial findings between infected patients for detecting temporal changes from February 2020. In a hospital-based open-cohort study, patients are followed from admission until one year from their discharge (the 1st, 4th, 12th weeks, and the first year). The patient recruitment phase finished at the end of August 2020, and the follow-up continues by the end of August 2021. The measurements included demographic, socio-economics, symptoms, health service diagnosis and treatment, contact history, and psychological variables. The signs improvement, death, length of stay in hospital were considered primary, and impaired pulmonary function and psychotic disorders were considered main secondary outcomes. Moreover, clinical symptoms and respiratory functions are being determined in such follow-ups. Among the first 600 COVID-19 cases, 490 patients with complete information (39% female; the average age of 57±15 years) were analyzed. Seven percent of these patients died. The three main leading causes of admission were: fever (77%), dry cough (73%), and fatigue (69%). The most prevalent comorbidities between COVID-19 patients were hypertension (35%), diabetes (28%), and ischemic heart disease (14%). The percentage of primary composite endpoints (PCEP), defined as death, the use of mechanical ventilation, or admission to an intensive care unit was 18%. The Cox proportional-hazards model for PCEP indicated the following significant risk factors: Oxygen saturation < 80% (HR= 6.3; [CI 95%: 2.5,15.5]), lymphopenia (HR= 3.5; [CI 95%: 2.2,5.5]), Oxygen saturation 80%-90% (HR= 2.5; [CI 95%: 1.1,5.8]), and thrombocytopenia (HR= 1.6; [CI 95%: 1.1,2.5]). This long-term prospective Cohort may support healthcare professionals in the management of resources following this pandemic.

## Introduction

The 2019 novel coronavirus disease (COVID-19) epidemic was officially announced by the World Health Organization (WHO) as an international public health emergency. Aggressive growth in the number of those affected with COVID-19 makes this virus such a threat. Patients were assessed for viral pneumonia through the ascertainment and testing of bronchoalveolar lavage fluid utilizing whole-genome sequencing, cell cultures, and polymerase chain reaction (PCR). In addition to high mortality rate, the disease has caused severe psychological problems among patients [1]. The clinical characteristics of COVID-19 patients were frequently reported [2]. Over 200 countries have a substantial incidence to date, including countries from the Middle East, North America, Asia, Australia, and Europe [3]. Currently, when COVID-19 is rapidly scattering worldwide, and the number of cases in the Middle East is rising with increasing pace in several affected areas, there is a need for immediate targeted action.

These high-risk procedures have implications for organization and medical practice of hospital care during this outbreak. Health policymakers everywhere plan for pandemics because their decisions can cause sharp shocks to societies and require a substantial and massive change in health system capacity [4]. These problems, caused by the COVID-19, emphasizes the importance of analyzing the epidemiological data worthwhile. The COVID19 studies have typically been focused on the initial clinical characteristics and the epidemiological description [5]. A majority of infected patients had mild to severe lung abnormalities on their chest CT scans when they were discharged from the hospital. Experts believe such groups may need closer follow-ups [6].

According to our understanding, most of the studies related to this outbreak identify the epidemiology and clinical characteristics of infected patients [7], the genomic classification of the virus [8], and trials for global health governance [9]. COVID-19 Cohorts in the literature are either retrospective or in a short duration (less than three months) [10–12]. However, to the best of our knowledge, there is no study examining the changing status of this virus and its psychological impact on the infected patients in the Middle East. Although the epidemic is still ongoing, initial lessons from its spread can help inform public health officials and medical practitioners to combat its progression. Accordingly, long-term prospective Cohorts are valuable studies in this pandemic.

Iran is one of the countries with a high prevalence of COVID-19. It has been revealed that half of the Iranians have limited health literacy, which is more common in exposed groups, such as unemployed people, homemakers, and older people [13]. Isfahan, the largest city in central Iran, with about 23000 positive corona cases, is among the three top Iranian cities with a high outbreak. For the improvement of future preparedness plans and provide a critical assessment of the resources and actionable items for stopping COVID-19 spread, utilizing lessons learned from the coronavirus outbreaks in hot zones could be helpful.

In this article, we present an effort to introduce an open-cohort from Isfahan, Khorshid COVID Cohort (KCC) study, to compile and analyze epidemiological outbreak information of COVID-19 infected patients. The objectives of our study are to provide a longitudinal overview of the patients’ condition and identify different related risk factors. This longitudinal study aimed to analyze the following signs and symptoms findings in patients with COVID-19 pneumonia for temporal changes and establish the incidence of psychological disorders and related prevalent symptoms after discharge from the hospital.

It is the first study where the temporal progression between infected patients in the Middle East is explored, to the best of our knowledge. Our research data are looked-for developing evidence-driven policies to reduce the adverse effects of the increased outbreak and psychological impacts. It may help government agencies and policy health makers protect the psychological well-being of society in the face of the COVID-19 outbreak in Iran and different parts of the world.

## Materials and methods

### Study population

In this prospective hospital-based surveillance study, patients admitted for COVID-19 from February 2020 until September 2020 in the Khorshid Hospital in Isfahan were recruited (Fig 1). Khorshid is the referral hospital for COVID-19 adults in Isfahan. About fifty percent of the entire COVID-19 population from Isfahan refers to this hospital.

**Fig 1.**
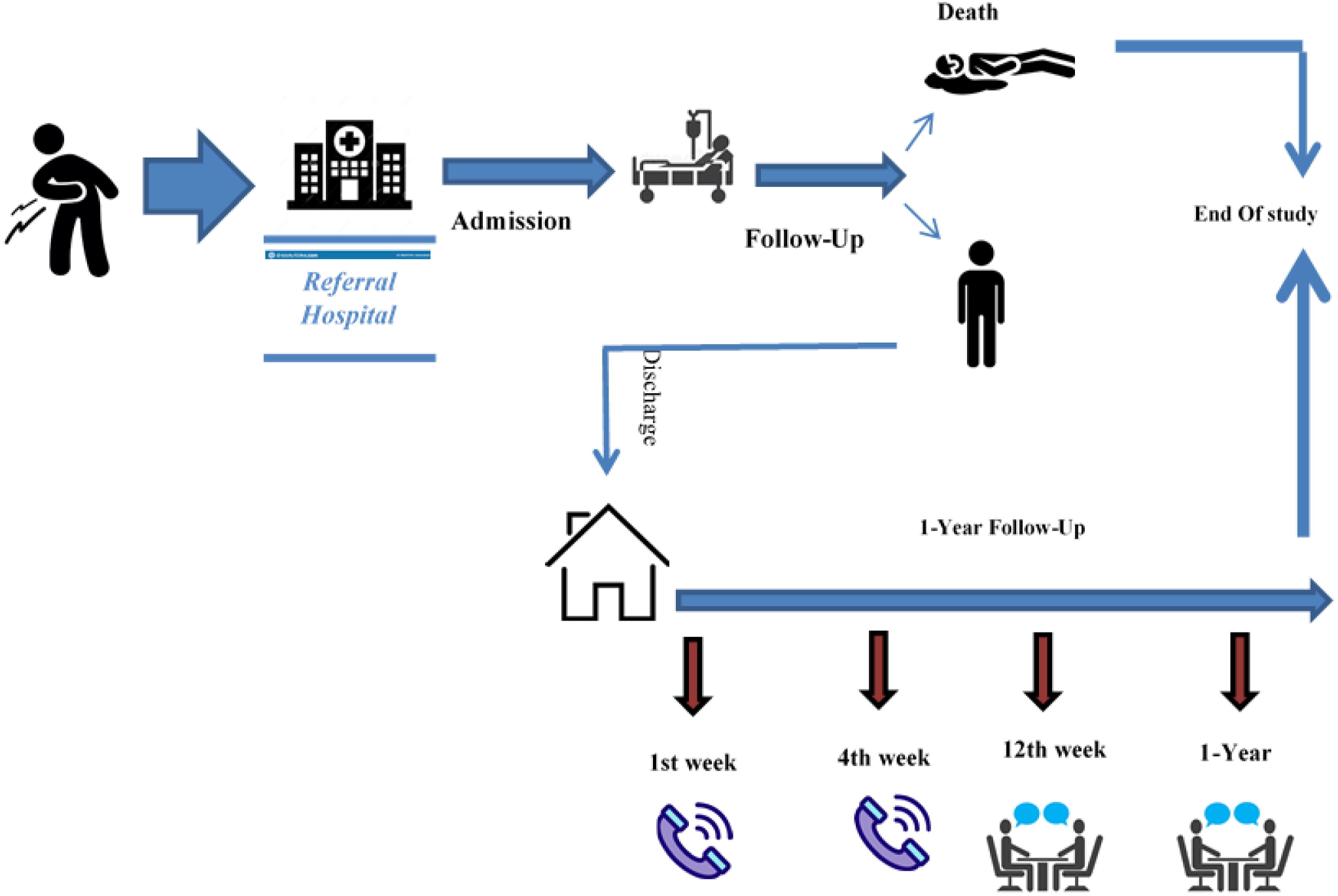
The scheme of the Khorshid COVID Cohort (KCC) study.

In this study, we diagnosed the COVID-19 patients based on the Chinese COVID-19 diagnosis and treatment guidelines, and the WHO provisional advice [14]. Then, in case of a positive polymerase chain reaction (PCR) or compatible Chest computed High-resolution CT (HRCT) scan with COVID-19, the patients were recruited in this study. The ethics committee of the Isfahan University of Medical Sciences (IUMS) and the other national authorities approved the study (IR.MUI.MED.REC.1399.029). The experimental protocol was also conformed to the Declaration of Helsinki. Also, the entire subjects gave written informed consent to the experimental procedure. The written informed consent was given by the first relative family of patients with severe conditions. No minors participated in our study. This Cohort has two phases. The first phase is on the admission information on hospitalized patients until discharge or death. In contrast, the second phase is related to patients discharged from the hospital for future symptoms or social factors. Six-hundred patients were enrolled in the first phase, while four-hundred ninety patients with complete information were analyzed in this paper.

### Baseline information collection

We obtained the information related to demographic, socioeconomic status (SES), medical history, underlying chronic diseases, chest computed tomographic (CT) scans, signs, symptoms, laboratory findings, treatment (including oxygen support, antibiotics, antiviral therapy, corticosteroid therapy) during the hospital admission, and outcome data from patients medical records by Table 1 checklist. The body temperatures of the patients were taken at least four times a day. The detection of nucleic acid from the SARS-CoV-2 in upper respiratory specimens was qualitatively assessed using the PCR test. The PCR was performed once for admission. Moreover, the Charlson Comorbidity Index (CCI) [15], an indicator of comorbidity, was calculated for each COVID-19 patient.

**Table 1.**
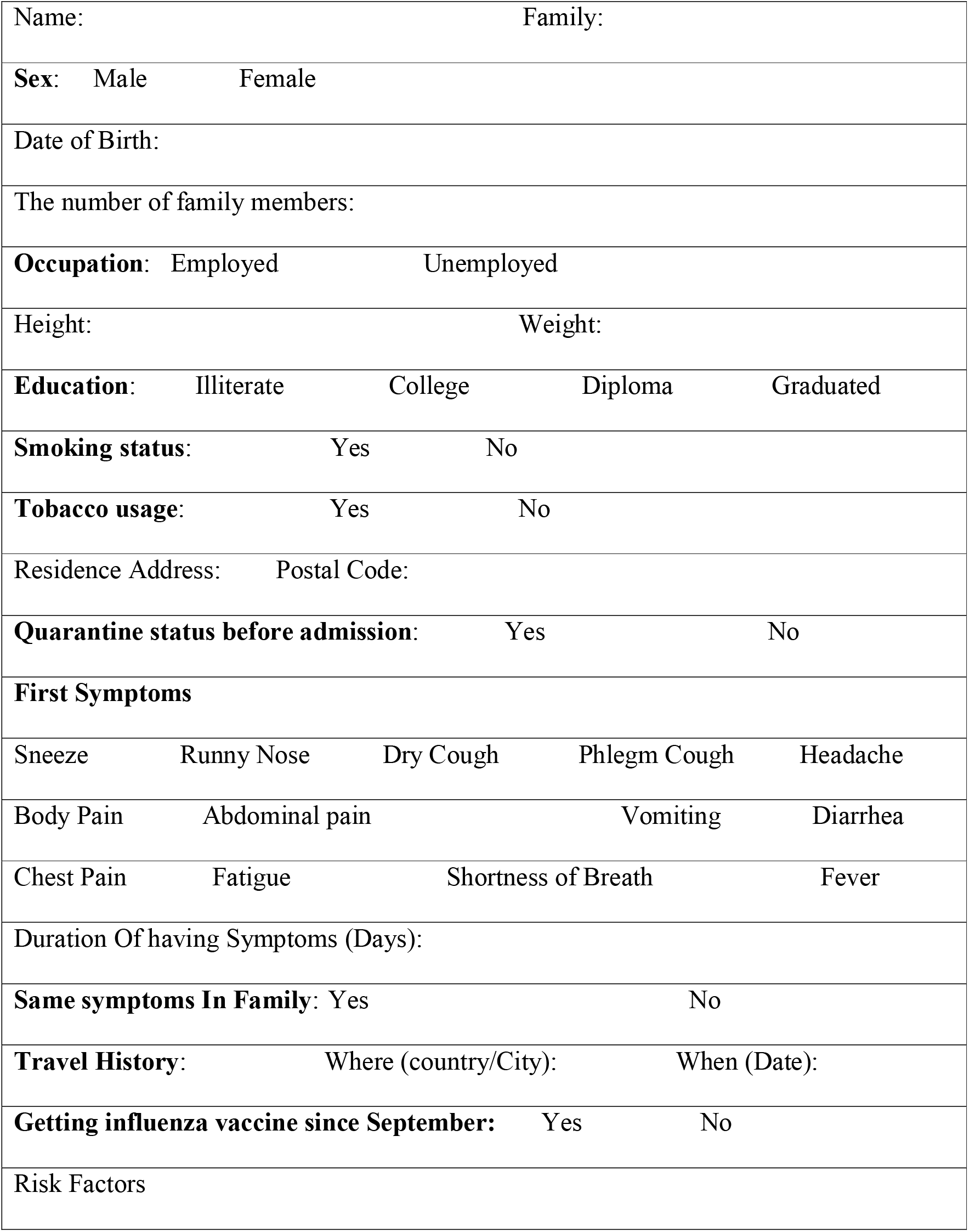
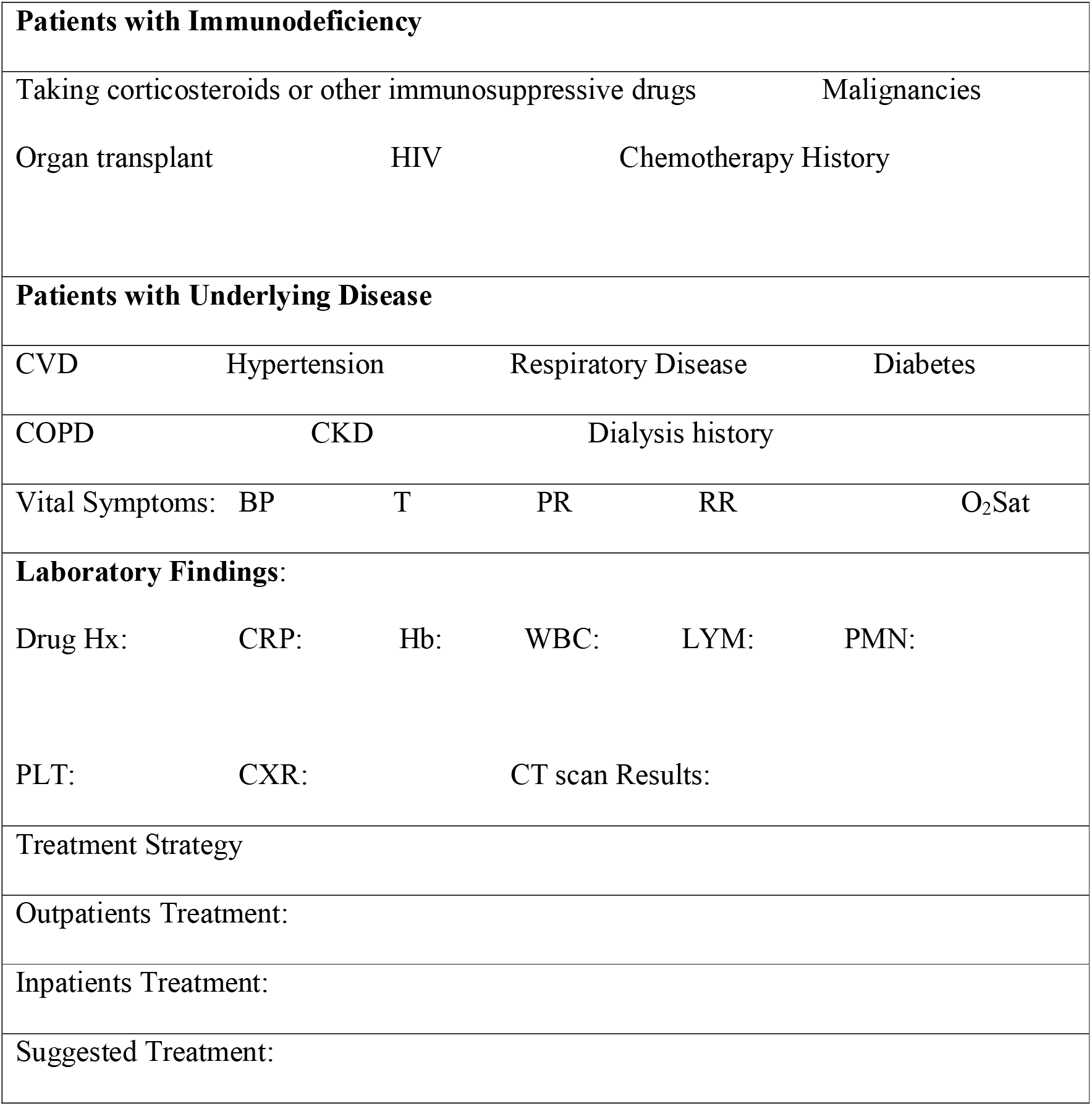
Checklist of following patients admitted to the hospital.

|  |  |  |  |  |
| --- | --- | --- | --- | --- |
| Name: |  | Family: |  |  |
| **Sex:** | Male | Female |  |  |
| Date of Birth: |  |  |  |  |
| The number of family members: |  |  |  |  |
| **Occupation:** | Employed | Unemployed |  |  |
| Height: |  | Weight: |  |  |
| **Education:** | Illiterate | College | Diploma | Graduated |
| **Smoking status:** | Yes | No |  |  |
| **Tobacco usage:** | Yes | No |  |  |
| Residence Address: |  | Postal Code: |  |  |
| **Quarantine status before admission:** |  | Yes | No |  |
| **First Symptoms** |  |  |  |  |
| Sneeze | Runny Nose | Dry Cough | Phlegm Cough | Headache |
| Body Pain | Abdominal pain | Vomiting |  | Diarrhea |
| Chest Pain | Fatigue | Shortness of Breath |  | Fever |
| Duration Of having Symptoms (Days): |  |  |  |  |
| **Same symptoms In Family:** |  | Yes | No |  |
| **Travel History:** | Where (country/City): |  | When (Date): |  |
| **Getting influenza vaccine since September:** |  | Yes | No |  |
| Risk Factors |  |  |  |  |

|  |  |  |  |  |  |
| --- | --- | --- | --- | --- | --- |
| **Patients with Immunodeficiency** |  |  |  |  |  |
| Taking corticosteroids or other immunosuppressive drugs |  |  | Malignancies |  |  |
| Organ transplant | HIV | Chemotherapy History |  |  |  |
| **Patients with Underlying Disease** |  |  |  |  |  |
| CVD | Hypertension | Respiratory Disease |  | Diabetes |  |
| COPD | CKD | Dialysis history |  |  |  |
| Vital Symptoms: | BP | T | PR | RR | O<sub>2</sub>Sat |
| **Laboratory Findings:** |  |  |  |  |  |
| Drug Hx: | CRP: | Hb: | WBC: | LYM: | PMN: |
| PLT: | CXR: | CT scan Results: |  |  |  |
| Treatment Strategy |  |  |  |  |  |
| Outpatients Treatment: |  |  |  |  |  |
| Inpatients Treatment: |  |  |  |  |  |
| Suggested Treatment: |  |  |  |  |  |

### Laboratory findings

Pharyngeal swab samples were collected for the COVID-19 test on arrangement. Blood samples were collected from each participant, and routine blood test, including Lymphocyte count (LYM), Platelet, Neutrophil (NEU), and White blood cell (WBC) counts, Hemoglobin, Sodium (Na), Potassium (K), Blood urea nitrogen (BUN), Blood creatinine (Cr), Blood Calcium (Ca), Potassium (P), and Magnesium (Mg) were performed on the blood samples. Furthermore, blood biochemistry parameters such as C-reactive protein (CRP), Aspartate aminotransferase (AST), Alkaline phosphatase (ALP), Alanine aminotransferase (ALT), Urea, as well as lactate dehydrogenase (LDH) and Albumin were assessed using HITACHI 7600-020 automated biochemistry analyzer. Other requested parameters on admission included troponin and EKG.

### Imaging protocol

We acquired the entire chest CT images using a 64-slice Philips scanner, with low-dose protocol, between patients in the supine position, without the injection of contrast, and at full inspiration. The images were evaluated by two radiologists with experience in thorax imaging. A senior investigator with at least ten years of experience at interpreting chest CT images resolved any disagreement. The following characteristics were recorded for each CT scan: distribution (craniocaudal, and transverse), the pattern, and disease severity according to a semi-quantitative scoring system [16]. Each CT scan was sub-classified according to published RSNA guidelines to the following four groups: typical, indeterminate, atypical, and negative [17].

### Discharge criteria

Patients presenting the following criteria were discharged: per evaluation of the treating physician, the clinical symptoms significantly improved (respiratory rate <20, pulse rate<100, oxygen saturation of 92% while the patient was breathing ambient air), AND the body temperature to be returned to normal for more than two days without any antipyretic medications, AND normal swallow for the solid oral medication (whole tablets and capsules), AND passing until 14 days after the onset of symptoms for patients without suitable caring system, AND good facilities for quarantine at home [18].

### Follow-up

The total follow-up time lasts for one year in our study (The 1st, 4th, 12th weeks, and the first year). The following information of the discharged patients are being recorded: clinical signs and symptoms at home, possible COVID-19 recurrence, underlying diseases in their family, psychological symptoms, sleep quality, and related problems (Table 2). Discharged patients are being followed up by telephone in the first and fourth weeks to obtain whether they had any symptoms presented in Table 2. In the 12th week and the first year, each patient attends the hospital to complete the Patient Health Questionnaire-9 (PHQ-9) and the Depression Anxiety Stress Scales (DASS-21) [19]. Moreover, clinical symptoms and the respiratory function (e.g., maximal inspiratory pressure (MIP), maximal expiratory pressure (MEP), and forced vital capacity (FVC), and forced expiratory volumeone second (FEV1)) are being determined in the last two follow-ups.

**Table 2.**
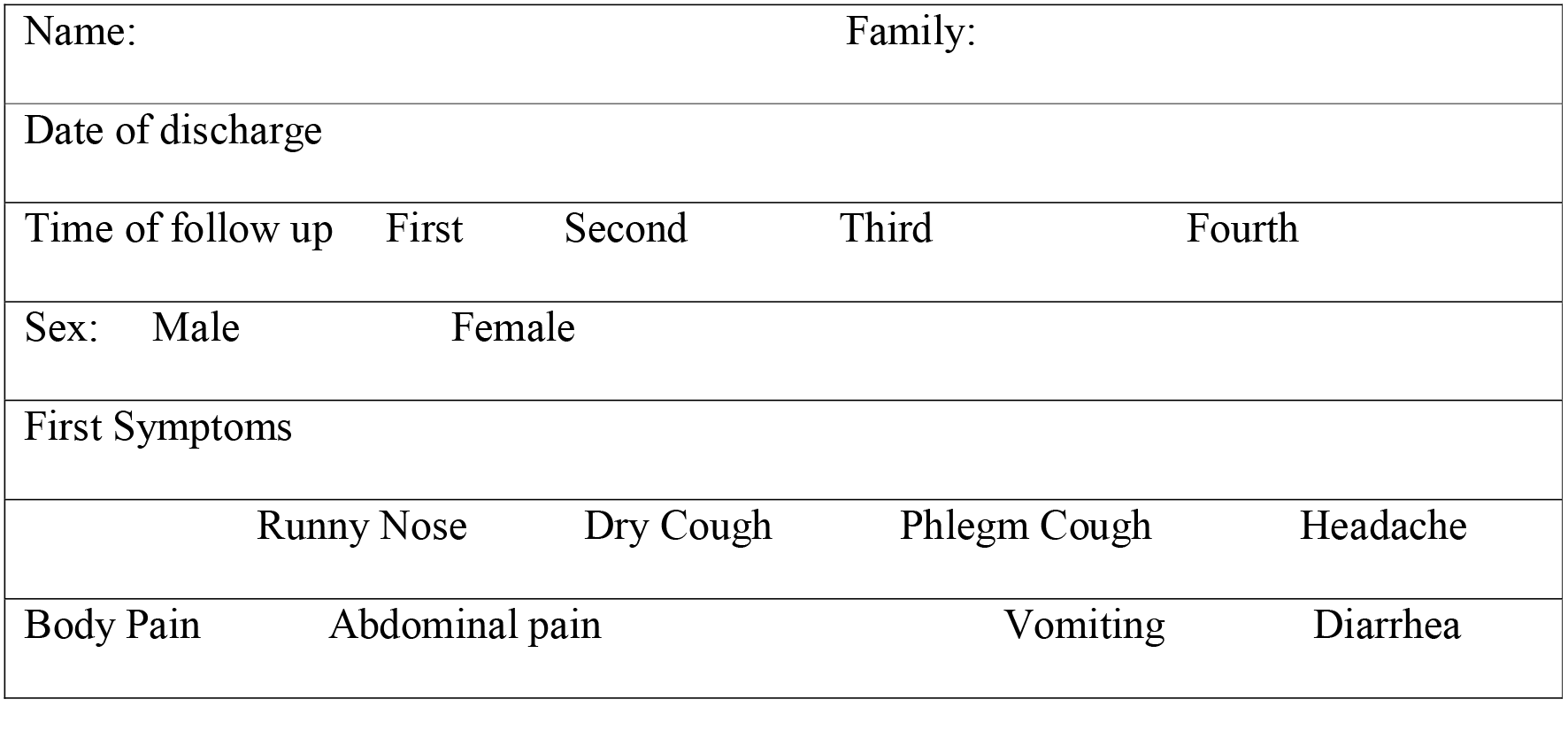
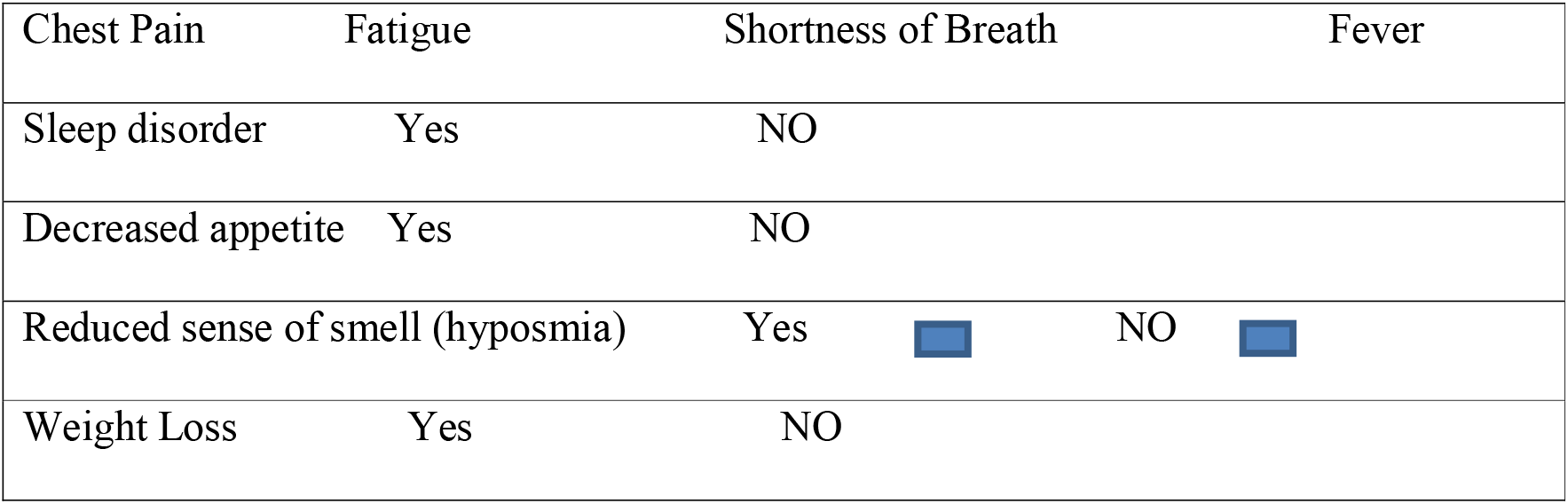
Checklist of following patients discharged from the hospital.

We used the self-report PHQ-9 questionnaire to measure the depression severity, with the total scores labeled as the following: severe depression (15–21), moderate depression (10–14), mild depression (5–9), and no depression (0–4) [20]. Depression, Anxiety, and Stress (DASS-21), a.k.a. Mental health status was measured based on previous research [21]. The stress subscale was assessed using question numbers one, six, eight, eleven, twelve, fourteen, and eighteen. The total stress subscale score was separated into five groups: extremely severe stress (35–42), severe stress (27–34), moderate stress (19–26), mild stress (11–18), and normal (0–10). The anxiety subscale was estimated using question numbers two, four, seven, nine, fifteen, nineteen, and twenty. The anxiety subscale score was also divided into five groups: extremely severe anxiety (20–42), severe anxiety (15–19), moderate anxiety (10–14), mild anxiety (7–9), and normal (0–6). The depression subscale was formed using questions three, five, ten, thirteen, sixteen, seventeen, and twenty-one. The total depression subscale score was classified as extremely severe depression (28–42), severe depression (21–27), moderate depression (13–20), mild depression (10–12), and normal (0–9). It was shown that the DASS is a reliable and valid method in measuring mental health between the Iranian population [22]. Note that the DASS was previously used in the literature for the other coronavirus version like SARS [23,24].

### Outcomes

The initial results for the admitted patients were the following: Vital signs improvement, including SpO2, pulse and respiratory rate, blood pressure, temperature, intubation rate, death, and length of stay at the hospital. The secondary outcomes are impaired pulmonary function, later signs and symptoms, psychotic disorders, sleep disorders, and sustained end-organ failure. Moreover, The primary composite endpoint (PCEP), defined as death, the use of mechanical ventilation, or admission to an intensive care unit (ICU) [25], is reported in this paper.

The research team of professionals from Khorshid Hospital and clinical faculty members of Isfahan University of medical sciences cross-checked the data. A trained team of researchers independently entered the data into a computer-based database. In a case of missing in the raw data, the coordinators were requested for clarification to contact the corresponding clinicians. We obtained data from their pre-admission information based on medical histories and through contact with their close relatives considering medical records from previous hospital visits for patients who had consciousness problems on admission. Further details of the KCC based on the STROBE statement were provided (S1 File). Such big data is being analyzed by our data mining and biostatistical teams.

### Data analysis

First, we entered the data into the Epi-Info 3.5.3 program (https://www.cdc.gov/epiinfo/). Then, the data were analyzed using STATA v12.0 (StataCorp, College Station, TX). The patient characteristics were reported as a percentage for categorical and mean (SD) for continuous data. The endpoint in this study was death or cure from the entire COVID-19-related causes. We confirmed the endpoint by reviewing hospital medical registration or by calling using the registered phone number. When the study period finishes, individuals alive after a follow-up time are censored. Accordingly, the subject outcome variable is death or censored after the follow-ups. In this manuscript, Hazard ratios and 95% confidence intervals for PECP associated with different baseline factors were estimated using Cox proportional-hazards regression models.

We use survival analysis to identify the connection between the patient’s attributes with time from initial admission to death or the end of the follow-up after discharge are considered as covariates. The life table is used to estimate survival after the first admission, and a log-rank test is used for survival curves comparison according to different events. Time-dependent Cox regression is used to calculate the adjusted hazard rate to determine independent predictors of time to death [26]. Statistical tests were two-sided, and P-values less than 0.05 were considered to indicate statistical significance.

### Results and discussion

Descriptive statistics were calculated for socioeconomic, demographic characteristics, clinical symptoms and health service utilization, contact history, and additional health information variables (Table 3). A total of 490 first COVID-19 cases with complete information were analyzed in this manuscript, with the average age of 56.58±15.09 years (39%, female) admitted to the Khorshid hospital from February 2020. Eight percent of the admitted patients were transferred to the intensive care unit (ICU) (48% male). Thirty-four patients (7%) died. The top three leading causes of admission were: fever (77%), dry cough (73%), and fatigue (69%). Sneeze (10%), runny nose (14%), and abdominal pain (17%) were the least frequent symptoms among COVID-19 patients. The top three prevalent comorbidities with COVID-19 were hypertension (35%), diabetes (28%), and ischemic heart disease (14%). The characteristics of 76 patients with pneumonia with a negative PCR test and CT-scan were provided in Table 3 as the control group. The clinical symptoms of the control and COVID-19 groups were similar, except for fatigue and dry cough. The control group was hospitalized due to the similar characteristics to COVID19 patients during the pandemic. The information of this group was provided instead of that of healthy subjects, as the discrimination between pneumonia and COVID-19 could be valuable.

**Table 3.**
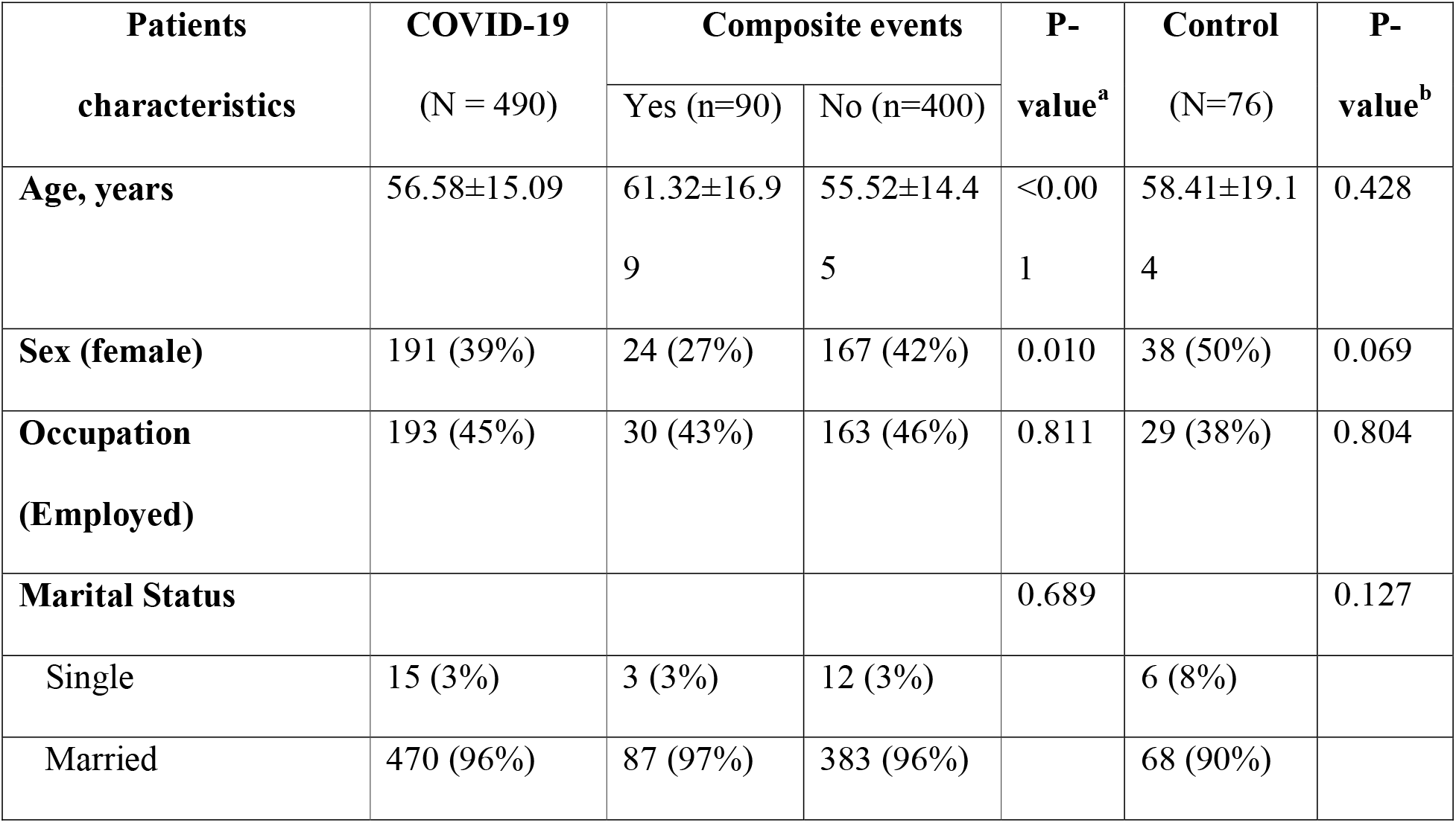
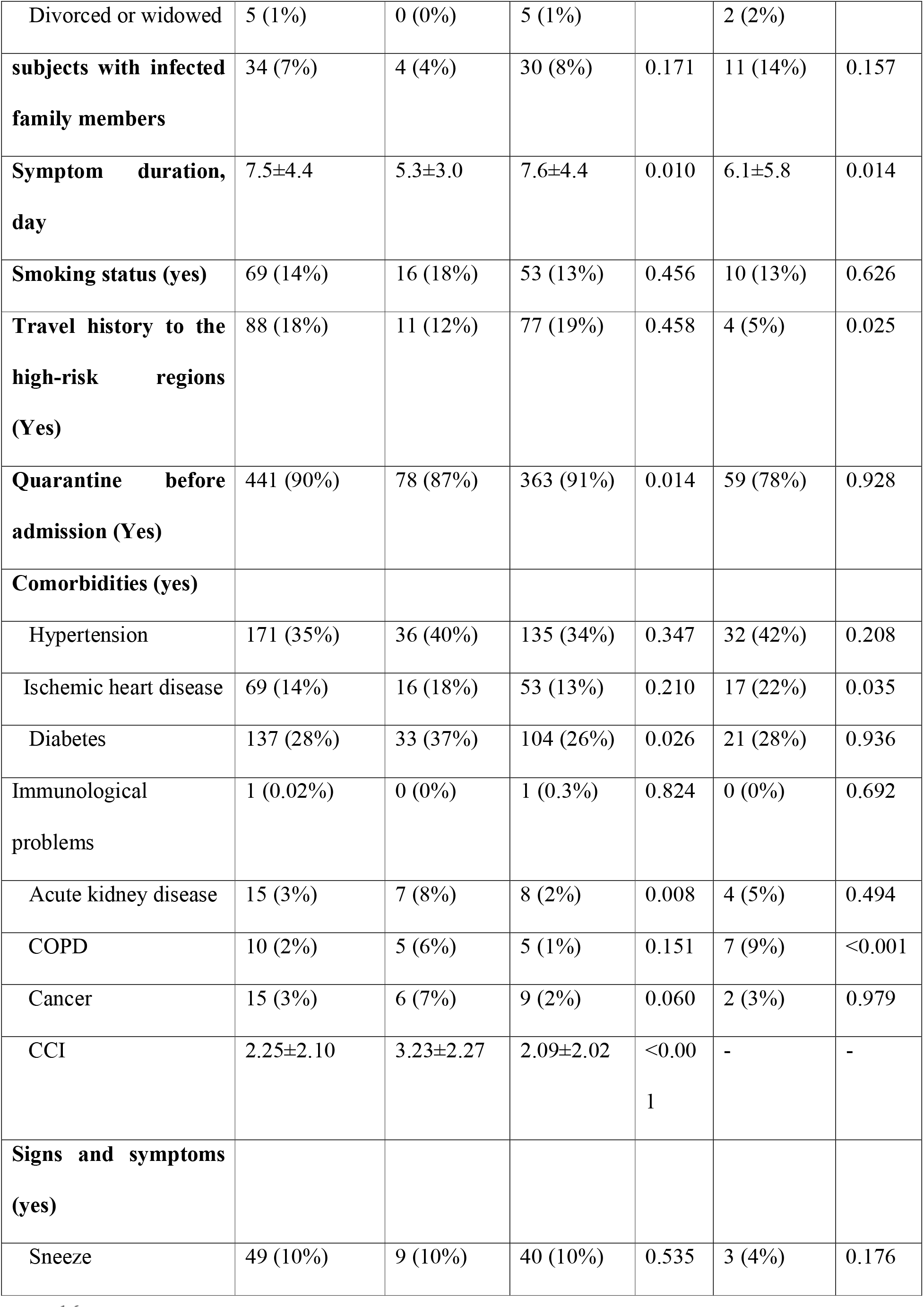
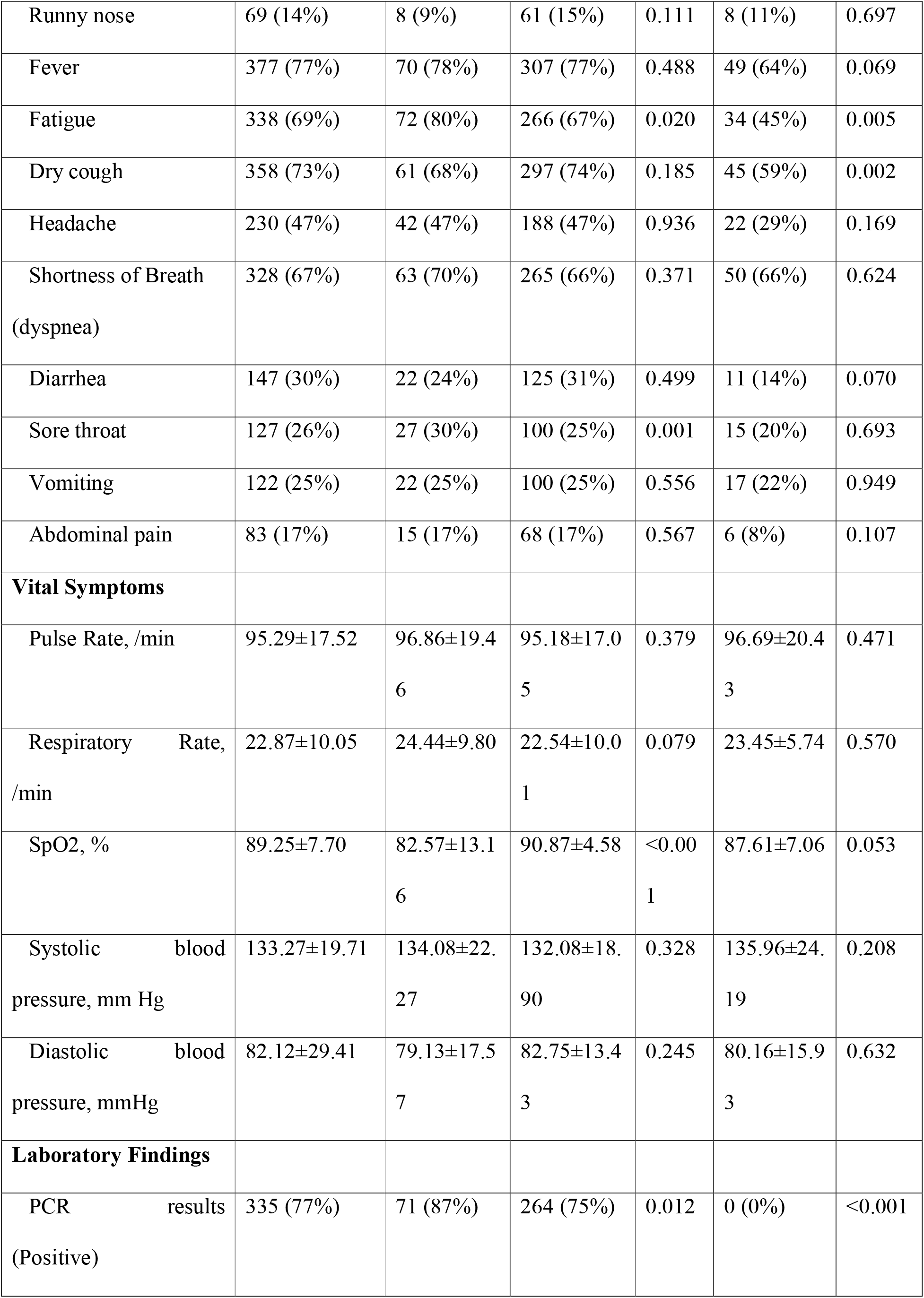
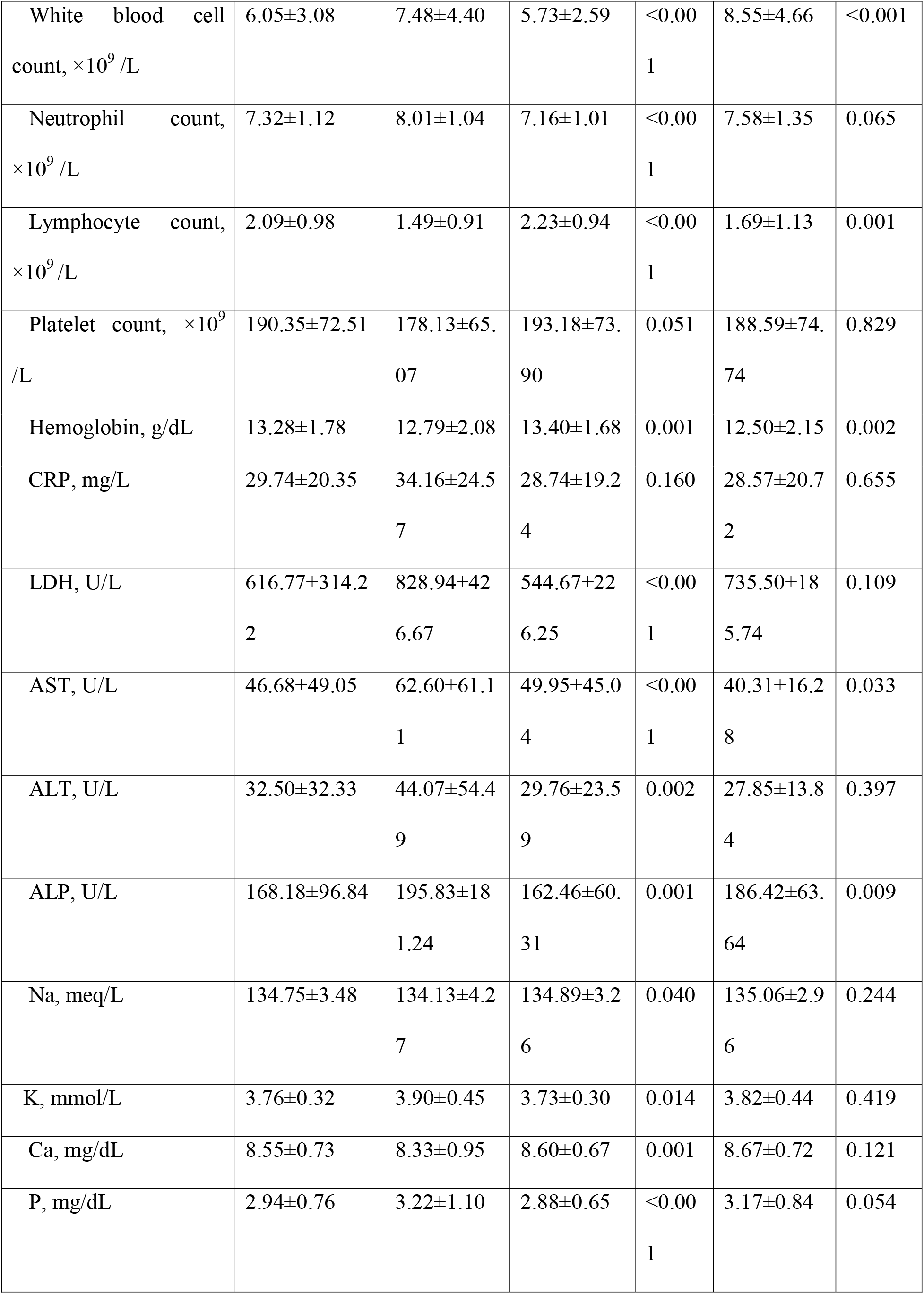
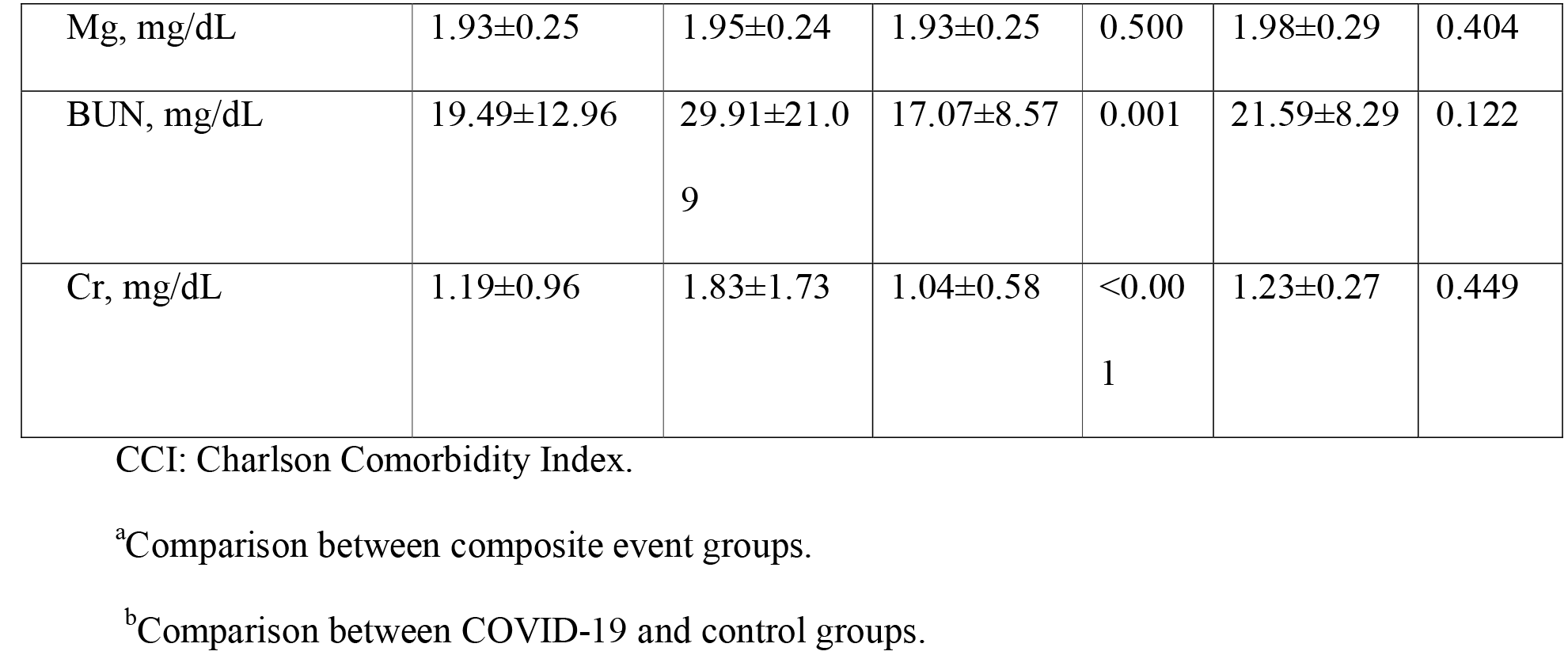
Demographic, SES, comorbidities, signs and symptoms, vital symptoms, and laboratory findings of 490 patients admitted to a COVID-19 referral hospital in Isfahan.

The Cox Proportional-Hazards Model was used for PCEP (Table 4). Age was categorized based on the Petrilli et al. study [27], and the reference category was set to 0-44 years old. The normal range of oxygen saturation (95%-100%) was set as the reference category in SpO2. The developed survival model showed very good diagnostic accuracy (AUC= 0.85) for discriminating COVID-19 patients with/without PECP.

**Table 4.** The Cox Proportional-Hazards Model for PCEP.

| Factor | HR | CI 95% | p-value |
| --- | --- | --- | --- |
| Sex (male) | 1.505 | [0.944,2.401] | 0.086 |
| Age cat0 | ref |  |  |
| Age cat1 | 1.278 | [0.529,3.087] | 0.586 |
| Age cat2 | 1.295 | [0.576,2.909] | 0.532 |
| Age cat3 | 2.077 | [0.925,4.666] | 0.077 |
| Age cat4 | 1.512 | [0.662,3.453] | 0.327 |
| SpO2 cat0 | ref |  |  |
| SpO2 cat1 | 0.674 | [0.270,1.681] | 0.397 |
| SpO2 cat2 | 2.495 | [1.075,5.793] | **0.033** |
| SpO2 cat3 | 6.275 | [2.537,15.524] | **<0.001** |
| lymphopenia | 3.457 | [2.189,5.457] | **<0.001** |
| thrombocytopenia | 1.598 | [1.042,2.449] | **0.0315** |
| AUC | 0.85 | [0.78,0.91] |  |

Age categories were 0-44, 45-54, 55-64, 65-74, $\geq 75$ years (cat0-cat4); SpO2 categories were $<80\%$ , 80%-89%, 90%-94%, 95%-100% (cat3-cat0); Lymphopenia: lymphocyte count $< 109 /L$ ; Thrombocytopenia: Platelet count $< 150 \times 109 /L$ ; AUC: area under the ROC.

Among 456 COVID-19 patients discharged from the hospital, three patients died during the first follow up (i.e., the first week) while one more patient died at the second follow up (the fourth week). Those patients who died were from the PCEP group. The clinical symptoms of the patients during four weeks of follow up are shown in Table 5.

**Table 5.**
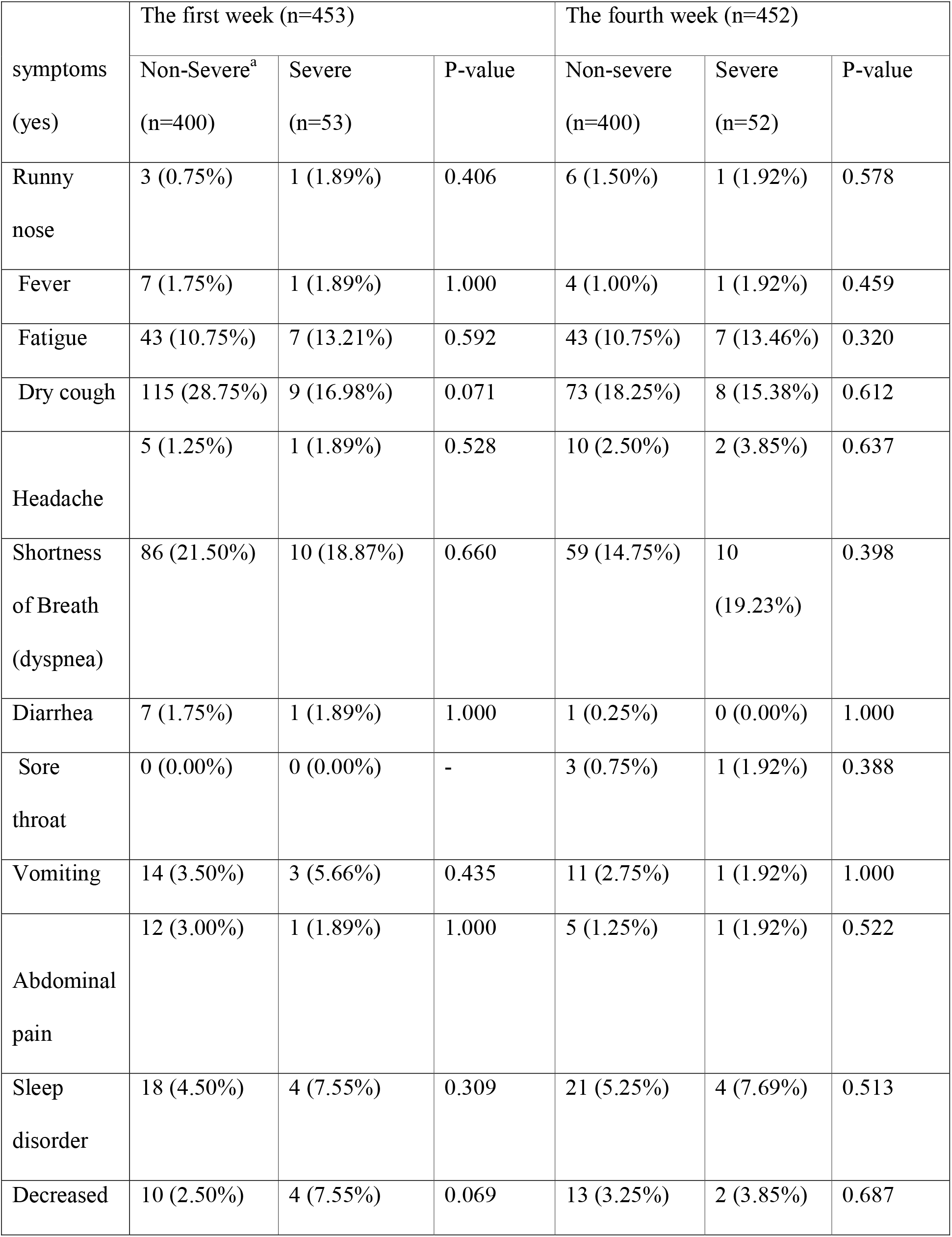
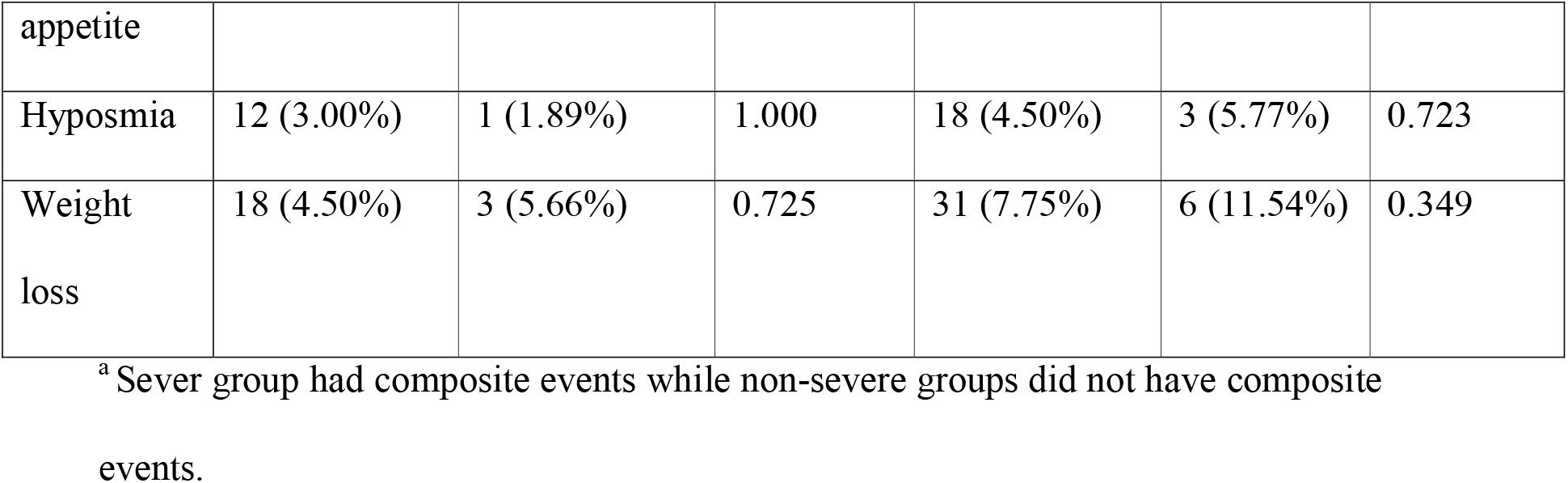
The clinical symptoms of the COVID-19 patients during four weeks of follow-up.

Among 490 COVID-19 patients, 90 patients had positive PCEP. The following characteristics were not significantly different in PCEP positive and negative groups: BMI, occupation, marital status, number of infected family members, smoking status, and travel history to the high-risk regions. However, the distribution of age (p<0.001), sex (p=0.010), symptom duration (p=0.010), and quarantine before admission (p=0.0140) were significantly different in PCEP positive and negative groups. Having analyzed the comorbidities in PCEP positive and negative groups, although hypertension, Ischemic heart disease, Immunological problems, COPD, and cancer were not significant, diabetes (p=0.026), and acute kidney disease (p=0.008) were significant.

Among signs and vital symptoms, only SpO2 (p<0.001), fatigue (p=0.020), and Sore throat (p=0.001) were significantly different in both groups. However, except for Mg (p=0.500), platelet count (p=0.051), and CRP (p=0.160), other laboratory findings were significant in the analyzed groups. Notably, elevated white blood cells (p<0.001), and neutrophil counts (p<0.001), BUN (p=0.001), LDH (p<0.001), AST (p<0.001), ALT (p=0.002), ALP (p=0.001), Cr (p<0.001), and K (p=0.014) were seen in PCEP positive groups compared with the negative groups. However, the platelet counts decreased in the PCEP group, but it was not significant (p=0.051). Moreover, the lymphocyte counts significantly decreased in the PCEP positive class (p<0.001).

The median time from admission to discharge was 5.0 days (interquartile range 3.0-8.0). The transmission route was by a history of exposure to the epidemic area (16%) or close contact with family members (25%), among which 10% of patients had both exposures. The average symptom duration was 7.5 days. A significant reduction of this time was observed in PCEP positive group respect to the negative group (p=0.010). Representative samples of the COVID-19 patients were provided at https://figshare.com/s/ef91187fb69a051b8396 (50 subjects with negative PCEP and ten subjects with positive PCEP).

The significant factors for PCEP survival analysis were Oxygen saturation < 80% (HR=6.275), lymphopenia (HR=3.457), Oxygen saturation (80%-90%) (HR=2.495), and thrombocytopenia (HR=1.598). It was found that hypoxemia is associated with in-hospital mortality and the oxygen saturation levels below 90% have high risks [28], which is in agreement with our results (Table 4). Lymphopenia is also associated with the disease severity of COVID-19 [29,30]. Moreover, thrombocytopenia was shown to be associated with the severity and mortality of COVID-19 [31,32]. Although not significant, older age is associated with mortality [33], similar to our findings. In our study, the PCEP risk was higher in men than women (HR=1.505; Tables 3 and 4), which is similar to Pérez-López et al. [34].

The clinical symptoms of the COVID-19 subjects discharged from the hospital were shown in Table 5 for the first- and fourth-week follow-ups. Such symptoms were similar in severe and non-severe cases, and the top three symptoms in the entire follow-ups and server or non-sever groups were dry cough, dyspnea, and fatigue, among which fatigue and cough were also observed in the three-month follow-ups of the study performed by Zhao et al. [35]. Moreover, the prevalence of sleep disorder, hyposmia, and weight loss increased from the first- to the fourth-week follow-ups (Table 5), emphasizing the long-term effect of COVID-19.

This Cohort aims to identify the problems presented in COVID-19 patients from first-time admission until death or one year after discharge. It is the first study in which COVID-19 patients are followed up to identify trends in signs and symptoms in an outbreak hot zone in the Middle East, to the best of our knowledge. It is required to perform thorough research on the novel, appearing in human infectious coronaviruses. It is then possible to explain their pathogenic mechanisms route and to recognize potential medicine. When considering the social effects of the outbreak, it might be possible to develop useful preventive and therapeutic medicaments. Moreover, the long-term analysis of psychological distress and mental illness symptoms of such patients after hospital discharge was taken into account in our Cohort. Such issues are fundamental, in general, in this pandemic [36].

According to our laboratory-confirmed cases, the common clinical manifestations were fever (77%), dry cough (73%), fatigue (69%), and shortness of breath (67%). The three most common clinical manifestations were consistent with the studies in China [7,25,37,38]. In laboratory examination results, less than 20% of the patients had decreased white blood cell counts. Also, the percentage of lymphocytopenia was 4%. It was in contrast with other studies that reported that most of their patients had this problem [25,39].

Previous studies demonstrated that the elderly and those with underlying disorders (i.e., cardiovascular diseases, diabetes, hypertension, and chronic obstructive pulmonary disease) developed rapidly into ARDS, even leading to ICU admission or death [7]. Although our findings presented that the death and ICU admission rate was 7% and 14%, a similar condition was observed. Cardiac complications, including heart failure, arrhythmia, or myocardial infarction, are common in patients with pneumonia.

It might be possible to reduce the probability of COVID-19 incidence and distress, reporting high-risk pathogens and social effects of this disease, which can help health policymakers lead reasonable policies when using our cohort results. Most people show mild symptoms. However, it may progress to ARDS, pneumonia, and multi-organ dysfunction in older people and those with comorbidities [40]. Many patients who survive acute viral pneumonia have impaired functions and health status in the first few months of recovery. However, longterm sequelae are still mostly unknown [41]. Patients who survived an episode of ARDS may have marked dyspnea and severe respiratory physiological abnormalities. Although a one-year post-ICU follow-up study showed that survivors of A(H1N1)-associated ARDS had minor lung disabilities [42], both obstructive and restrictive deficits have been reported in patients with ARDS. The ARDS survivors often suffer prolonged myopathy limiting daily living activities, post-traumatic stress disorder (PTSD), and an increased number of deaths after apparent recovery. Such problems usually exist for more than three months. Moreover, many patients show continuous neurocognitive dysfunction for one to two years. When managing these problems supportively, the knowledge of the PTSD persistency in such patients is required [43].

The medical and public health infrastructure and the economy were affected by this new virus outbreak worldwide [44]. Several studies have reported adverse psychological complications, including confusion, symptoms of traumatic stress, and anger in cases who encountered stressful situations. The long quarantine period, fear of disease, boredom, despair, inadequate food shortages, insufficient information, stigma, and financial loss are known as stressors in the COVID-19 epidemic [45,46]. Along with this, the primary purpose of this study is to identify psychological problems among discharged patients for one year.

It takes time to see how the virus affects our lives here in Iran and other parts of the world. It is also possible to have similar future outbreaks. Besides limiting this outbreak, preventive programs must be planned for such problems. It is of great importance when considering the warning by the WHO chief as “the virus will be with us for a long time.”

It must be mentioned that self-reporting used for the first two follow-ups has some limitations compared to face-to-face interviews for the second and third follow-ups. Also, psychological studies do not necessarily probe psychological services efficacy. Thus, dynamic observations and more follow-ups are necessary. The larger sample size is also required for result verification, which is available during the Cohort. Apart from such limitations, it is possible to perform a risk assessment and to use medical data mining for diagnosis and prognosis to identify complex interactions. Multistate models are the focus of our future work to provide the stacked probability plots and to predict the length of the hospital (or ICU) stay of COVID-19 patients.

## Conclusions

The COVID-19 outbreak has become a clinical danger to the general population and healthcare workers worldwide. However, knowledge about the trend of this novel virus, which leads to different symptoms and outcomes, remains limited. The practical option of different treatment, underlying disease, clinical findings, and different symptoms or signs on the disease prognosis is under evaluation and development. What we can do now is to design a cohort to follow patients to identify the underlying trend based on different outcomes and their risk assessment. It may assist healthcare policymakers and government agencies, and in the managing of health and resources following this pandemic.

## Data Availability

Representative samples of the COVID-19 patients were provided at https://figshare.com/s/ef91187fb69a051b8396 (50 subjects with negative PCEP and 10 subjects with positive PCEP).

## Acknowledgments

We would like to acknowledge the Khorshid hospital nurses and interns for this study, who recruited patients and collected follow-up data. Most importantly, we would like to acknowledge all the patients who consented to participate in this study. The authors are thankful to Prof. Martin Wolkewitz from the University of Freiburg for valuable comments.

## Supporting information

**S1 File. The specification of the KCC based on STROBE Statement**.

